# A 2SIR-VD Model for Optimizing Geographical COVID-19 Vaccine Distribution in the Philippines

**DOI:** 10.1101/2021.05.20.21257556

**Authors:** Allan Paolo L. Almajose, Angus White, Chelsea Diego, Red Lazaro, Nicanor Austriaco

## Abstract

COVID-19 is a novel respiratory disease first identified in Wuhan, China, that is caused by the novel coronavirus, SARS-CoV-2. It has triggered a global pandemic of historic proportions. The government of the Philippines began its national vaccine drive on March 1, 2021, with the goal of vaccinating seventy million of its citizens by the end of the calendar year. To determine the optimum geographical distribution strategy in the Philippines for the limited supply of vaccines that is currently available, we developed and adapted a basic SIR model that allows us to understand the evolution of a pandemic when public health authorities are vaccinating two susceptible populations within a country with different vaccine rates. Our analysis with our 2SIR-VD model revealed that prioritizing vaccine deployment to the National Capital Region (NCR) of the Philippines minimized the number of COVID-19 cases in the country. We therefore recommend deploying 90% of the available vaccine supply to the NCR to mitigate viral transmission there. The remaining 10% would allow the rest of the archipelago to vaccinate all of their senior citizens, thus shielding this vulnerable population against severe disease and death from COVID-19.

## INTRODUCTION

COVID-19 is a novel respiratory disease first identified in Wuhan, China, that is caused by the coronavirus, SARS-CoV-2 (Guan et al., 2020; Xie et al., 2020; Zhu et al., 2020). With widespread human-to-human transmission, the virus is highly contagious, and the COVID-19 pandemic is now of global concern (Burki, 2020; Paules et al., 2020).

On January 30, 2020, the Department of Health (DOH) of the Philippines reported its first case of COVID-19 in the country. The first case of local transmission was confirmed on March 7, 2020, with the first death due to local transmission reported on March 11, 2020. As of April 18, 2021, there have been 936,133 confirmed cases and 15,960 deaths from COVID-19 reported by the DOH throughout the archipelago (ncovtracker.doh.gov.ph).

The government of the Philippines began its national vaccine drive on March 1, 2021, with the goal of vaccinating seventy million of its citizens by the end of the calendar year (Inter-Agency Task Force for the Management of Emerging Infectious Dissease, 2021). The initial plan proposed to distribute vaccines equally across the geographic regions of the country according to priority groups by the government following WHO guidelines to mitigate mortality and morbidity. However, it is not clear if this geographic strategy of equal distribution across the archipelago would optimize the distribution of the limited numbers of available vaccines.

In this paper, we develop a basic SIR model that includes compartments for those who have been vaccinated (V) and those who have died (D) and adapt it to determine the optimum strategy to geographically distribute the limited stockpile of COVID-19 vaccines among the different regions of the Philippines. Though there have been other studies that have simulated the dynamics of a pandemic under different vaccine deployment conditions (Liu and Xia, 2011; Araz et al., 2012; Yu et al., 2016; Acuña-Zegarra et al., 2020; Goldenbogen et al., 2020; Yang et al., 2020; Cheng et al., 2021; Martínez-Rodríguez et al., 2021; Sjödin et al., 2021), we opted for a simpler model that would allow us to use the real time data published by the Department of Health of the Philippines, with all of its limitations and shortcomings.

Using our 2SIR-VD model, we compared five geographical distribution strategies to determine which one would most efficiently end the pandemic in the Philippines: 1) equal distribution throughout the country; 2) priority distribution to the NCR; 3) priority distribution to the “NCR Plus” bubble established by the Philippine government during the 1Q of 2021; 4) priority distribution to the three major metropolitan areas of Metro Manila, Metro Cebu, and Metro Davao; and 5) priority distribution to the NCR Plus, Metro Cebu, and Metro Davao. Our analysis revealed that prioritizing vaccine deployment to the NCR minimized the number of COVID-19 cases in the country. We therefore recommend first deploying 90% of the available vaccine supply to the NCR to mitigate viral transmission there. The remaining 10% would allow the rest of the archipelago to vaccinate all of their senior citizens, thus shielding this vulnerable population against severe disease and death from COVID-19.

## MODELLING METHODS

### Epidemic Modeling

A standard susceptible-infected-recovered (SIR) compartmental model was modified to include vaccinated (V) and deceased (D) populations in order to study the effects of alternative vaccination strategies on the evolution of a pandemic. In deriving the compartmental diagram and models, the following assumptions were made:

1. Population rates for infection recovery and death are assumed to follow the standard first order behavior with respect to the infected population;
2. Infection rates are controlled by a standard autocatalytic first-order behavior;
3. Vaccination rates are held as constant, that is, the number of vaccinations per unit time is not dependent to the amount of currently susceptible to infection, or vaccinated individuals;
4. Recovered individuals are no longer susceptible to reinfection; and
5. Vaccination is assumed to be 100% effective, that is, vaccinated individuals are no longer susceptible to reinfection.

We are aware that these assumptions are idealizations that will prevent us from making real time predictions for the length of a particular pandemic as vaccination rates are varied. However, they will allow us to accomplish the principal goal of this analysis, which is to compare the relative efficiencies of vaccination strategies that prioritize different geographical regions in the Philippines.

It is common practice to express epidemic modelling assumptions in terms of a compartmental model diagram. Diagram (1) shows the susceptible-infected-recovered-vaccinated-deceased (SIR-VD) compartmental model diagram used to simulate the behavior of COVID-19 cases in the Philippines.

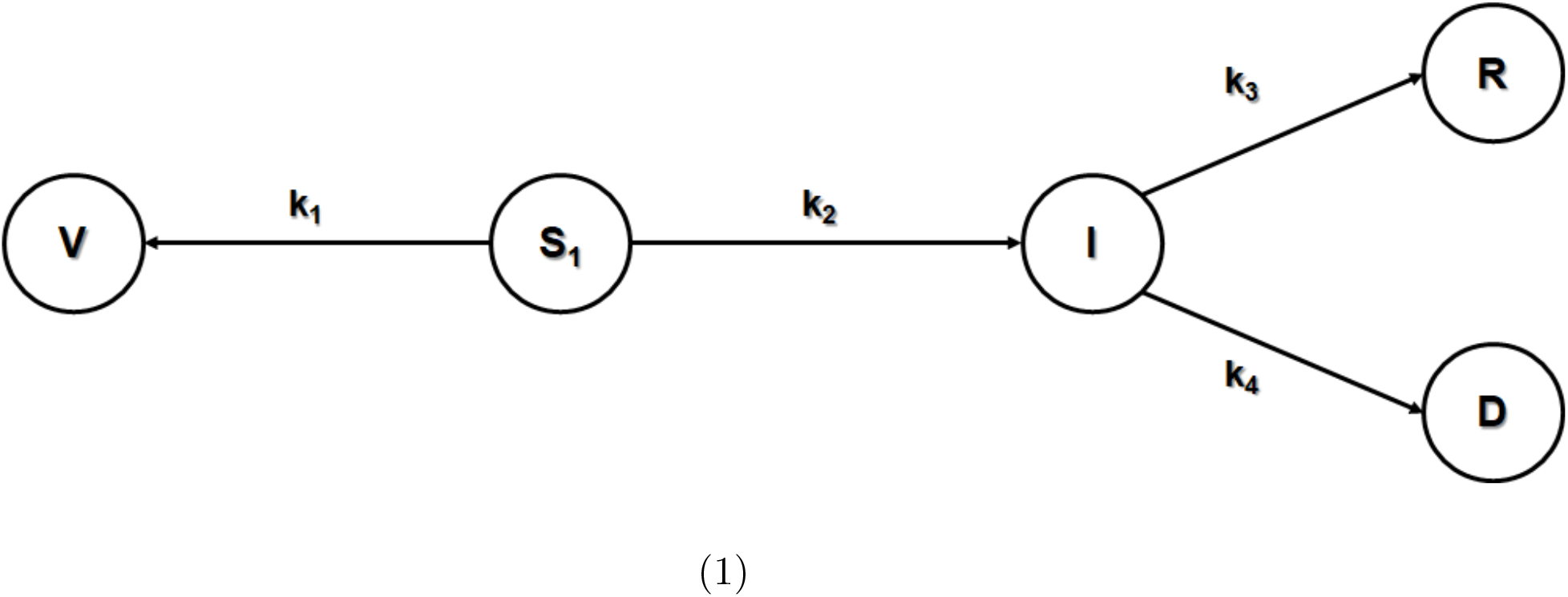

Expressed mathematically, equations (1) to (5) are the system of differential equations that describe the dynamic behavior of the infection.

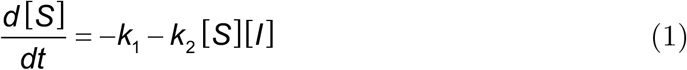

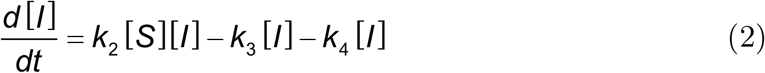

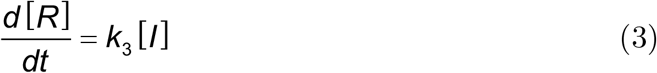

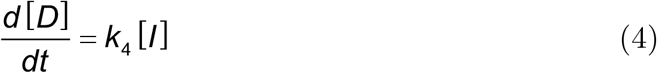

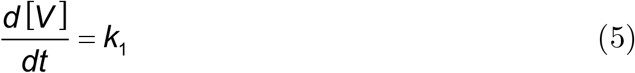

Constants *k*_2_, *k*_3_, and *k*_4_ describe the first-order infection, recovery, and death rate coefficients, which are normally found via regression of available datasets. Of utmost importance, however, is *k*_1_, as this is the coefficient that describes the vaccination rate of the susceptible population. Varying the values of *k*_1_ allows us to visualize the epidemic as it changes while the susceptible population is being inoculated. As a representation, assuming that the temporal dimension is in weeks, a dimensional analysis of equation (1) will directly show that the units of *k*_1_ is in number of persons at a given time;

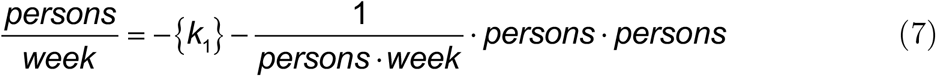

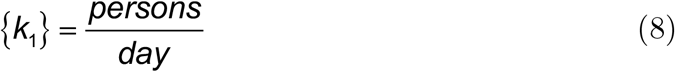

If population ratios are used instead of actual populations in the state variables of the model, as will be done in this paper, the rate *k*_1_ is simply the inverse of days:

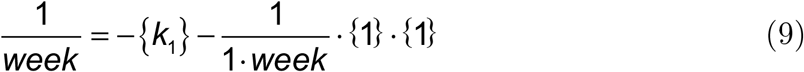

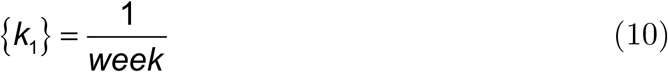

Should it be required to vaccinate a certain amount of susceptible individuals, *S*_*V*_, the value of *k*_1_ is calculated by dividing *S*_*V*_ with the total population of the system, *N*_*T*_ :

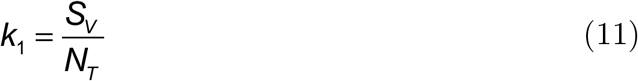

This SIR-VD model will allow us to understand how a pandemic evolves as a vaccination drive advances in a particular population.

We have also adapted our SIR-VD model to understand the evolution of a pandemic when public health authorities are vaccinating two susceptible populations with different vaccine rates. For simplicity’s sake, we assume that the infected individuals that belong to the distinct populations are indistinguishable, that is, there is an ideal mixing between the two infected populations. Diagram (2) graphically represents our two-population 2SIR-VD model with the underlying assumption that the two infected populations resulting from the susceptible individuals ideally mix.

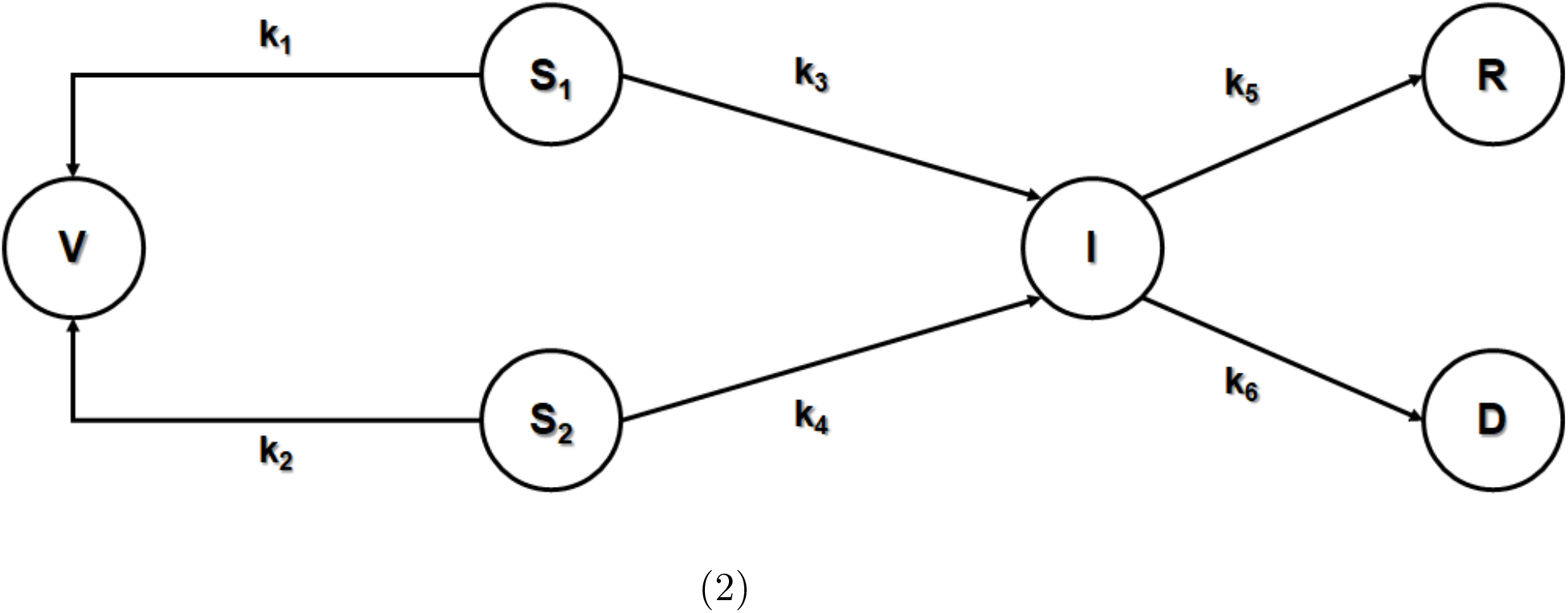

The model is mathematically represented by equations (12) to (17):

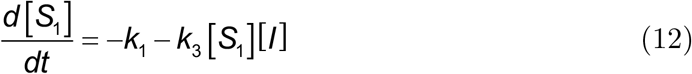

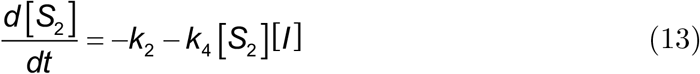

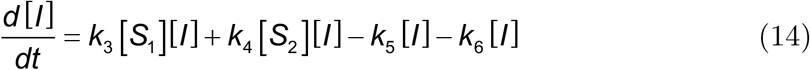

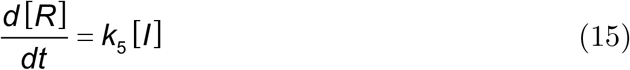

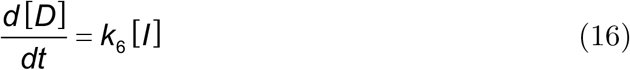

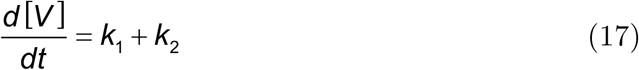

For equations (12) to (17), *k*_1_ describes the vaccination rate for *S*_1_ and *k*_2_ characterizes the vaccination rate for *S*_2_. Constants *k*_3_ to *k*_6_ describe the *S*_1_ infection rate, *S*_2_ infection rate, infection recovery rate and death rate.

Assuming that vaccination is limited to a certain amount *k*_*vac*_, the following equation must hold true:

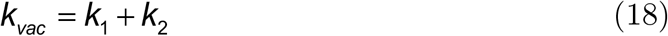

An important ramification of (18) is that the vaccinations may be weighted among the two susceptible populations. This allows us to study the progress of the epidemic as two populations are vaccinated with different rates. In this study, *S*_1_ will be designated as the target susceptible individuals, *S*_2_ will be assigned as the remainder of the total population of susceptible individuals. For example, in one of the scenarios we analyze in this paper, the population of Filipinos living in the National Capital Region (NCR) of the Philippines is designated *S*_1_, while the rest of the Filipino population living outside the NCR would be *S*_2_. In this way, *k*_1_ is set as the rate of vaccination of the target susceptible individuals which are normally less than the remainder of the total susceptible individuals. Should *S*_1_ evaluate to zero, the vaccination rate will be transferred to the remainder of the total susceptible individuals *S*_2_, that is:

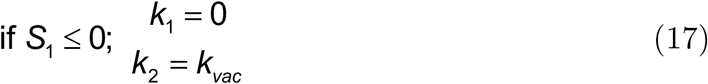

In principle, this 2SIR-VD model can also be used to evaluate the effect of vaccinating any two distinct populations within a certain geographical area that differ by age, by gender, or by residential location.

### Data Gathering and Numerical Solution Methods

COVID-19 data was extracted manually from the Philippine Department of Health’s (DOH) nCoV-2019 Tracker (ncovtracker.doh.gov.ph). The data used for this paper was preprocessed by evaluating the number of active cases, the cumulative recoveries, and cumulative deaths due to the epidemic per week as provided by the tracker. We chose to include epidemic information on a weekly basis to reduce the amount of data scatter and errors that we had observed because of delayed reporting of information by the DOH. Further, we discovered that this allowed us to properly account for the weekly recovery rates announced by the DOH.

In training the model, the weekly information between February 18, 2021 and March 31, 2021 was used. Given the disproportionate impact of the second surge on the overall numbers of the pandemic in the Philippines, we decided to limit our model to the 2021 first quarter surge of COVID-19 cases.

The numerical population values used in the study were taken from the Philippine Statistics Authority’s (PSA) QuickStat spreadsheets. A logistic model was used to predict the 2021 population values using the available 2000, 2007, 2010, and 2015 population information. The population numbers are displayed in Table 1.

**Table 1:** Population Values used for our Model Simulations. The numerical population values used in the study were taken from the Philippine Statistics Authority’s (PSA) QuickStat spreadsheets. A logistic model was used to predict the 2021 population values using the available 2000, 2007, 2010, and 2015 population information.

| **Geographical Region** | **Estimated**<br>**Population**<br>**(2021)** |
| --- | --- |
| Bulacan | 3,555,314 |
| Laguna | 3,425,983 |
| Rizal | 3,245,892 |
| Cavite | 4,227,720 |
| Metro Cebu | 3,430,503 |
| Metro Davao | 1,872,075 |
| NCR | 13,802,153 |
| Philippines | 112,868,636 |

In order to determine the rate for the systems of differential equations described by the 2SIR-VD model, numerical integration of the differential equations had to be applied with initial trial values for *k*_2_ to *k*_4_ using the standard SIR-V model, and trial values for *k*_3_ to *k*_6_ using the 2SIR-VD model. The fourth-order Runge-Kutta technique was used in integrating the systems of differential equations implemented in Microsoft Excel, with small temporal steps undertaken to assure numerical stability. Errors were calculated by evaluating the population ratio difference between those predicted by the mathematical models and the data. The normalized sum of squares of residuals is used as the objective function to train the rate constant data:

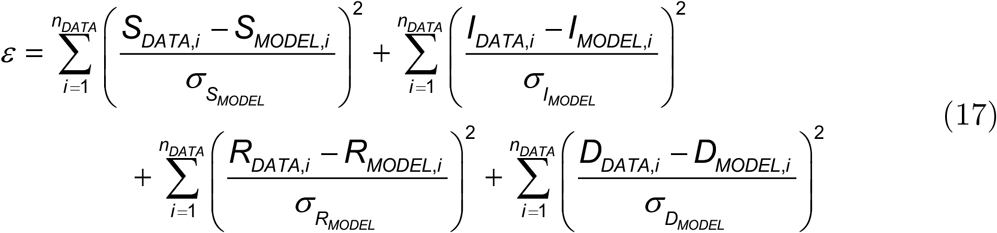

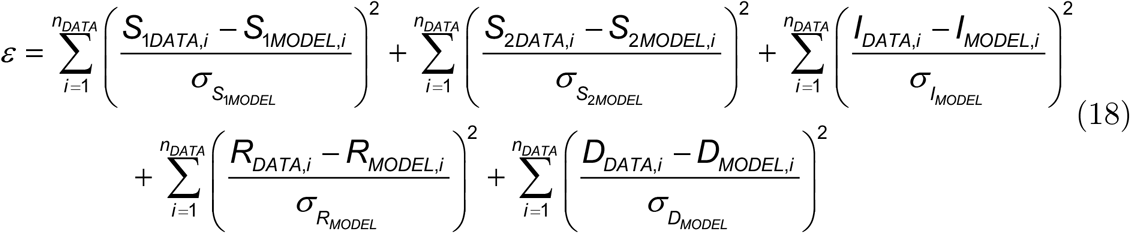

Equation (17) is applicable to the original SIR-VD model, while equation (18) was used to train the rate coefficients of the 2SIR-VD model.

To determine the final and converged rate constant values, the in-house Microsoft Excel Solver add-in’s Nonlinear Generalized Reduced Gradient method was used. Non-negativity constraints are assigned to the rate constants to facilitate convergence and correct behavior of the model. Further, central finite formulations have been used to evaluate the numerical derivative for the gradient, with the method converging at the error tolerance of 1×10^−8^.

An excel file that incorporates the 2SIR-VD model described here has been archived at GitHub with the following DOI: 10.5281/zenodo.475642.

## RESULTS & DISCUSSION

To compare potential COVID-19 vaccination strategies that prioritized different geographic populations in the Philippines, we developed a 2SIR-VD model that would allow us to simulate the spread of the pandemic while two different populations were being vaccinated at different rates. Because of the shortcomings and limitations of the available COVID-19 pandemic data provided by the Department of Health of the Philippines, we decided to construct a basic compartmental model with the minimum number of compartments that would allow us to test different vaccine deployment strategies that prioritized different geographical regions of the country.

To evaluate the quality and forecasting potential of our 2SIR-VD model, we began by comparing simulations of the pandemic at the beginning of the second surge of COVID-19 in the Philippines to the data made available by the Department of Health (DOH). As shown in Figure 1, our model accurately simulated the growth of the pandemic in the country as well as in the National Capital Region (NCR). Since the second surge of the pandemic was concentrated in the NCR, it is not surprising that the country-wide curve and the capital-only curve were similar in form and shape.

**Figure 1:**
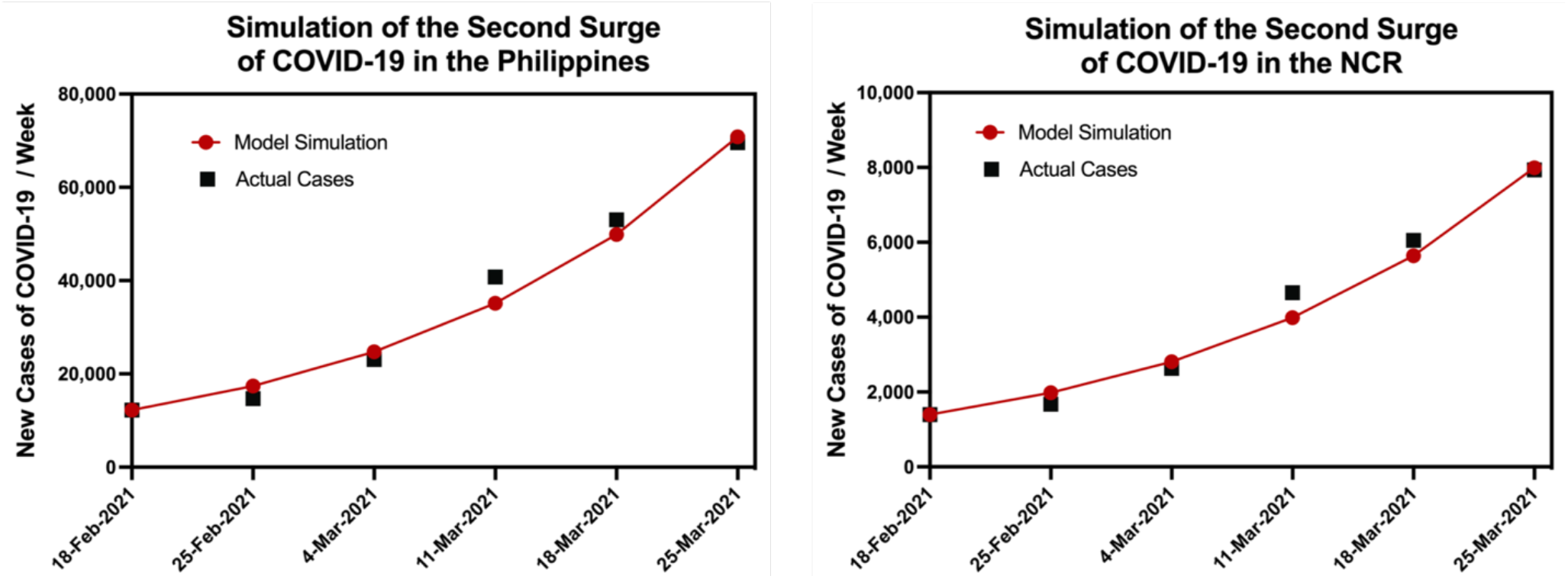
Simulations of the Second Surge of COVID-19 in the Philippines Compared to Real-time Data. Forecasted estimates of weekly new cases of COVID-19 for the Philippines and the National Capital Region compared to the real-time data provided by the Department of Health

Once we had determined that our model accurately simulated the growth of the pandemic during the second surge of COVID-19 in the Philippines, we used it to interrogate the impact of vaccinations on the dynamics of the pandemic. We chose to model the impact of a single-dose vaccine that conferred full protection against COVID-19. This is clearly an idealized scenario for the COVID-19 pandemic where most of the available vaccines require two doses separated by different periods of time, with different reported efficacy rates, but we believe that the model is sufficient to give public policy makers a sense of the impact of vaccination on the dynamics of the pandemic.

As shown in Figure 2, a simulation of the spread of the pandemic in the Philippines after the second surge of COVID-19 without any nonpharmaceutical interventions (NPI) like masking, social distancing, or lockdowns in place revealed that the surge would have peaked when the number of active cases of COVID-19 representing the individuals in the infected (I) compartment reached 11,345K. In contrast, the deployment of COVID-19 vaccinations at weekly rates of 500K, 1M, 2M, and 3M, would lower the peak to 73%, 49%, 18%, and 8% of its no vaccination value. Once again, we acknowledge that this is an idealized model that assumes equal distribution of the vaccination throughout the Philippines without any nonpharmaceutical interventions to halt the spread of the infection. The simulation is an attempt to discern the relative impact of vaccinations on the total number of COVID-19 cases in the country. The model predicts that the mathematical relationship between the weekly rate of vaccination and the decrease in COVID-19 cases is not linear. Rather, as vaccination rates rise, the comparable fall in total number of COVID-19 cases decreases.

**Figure 2:**
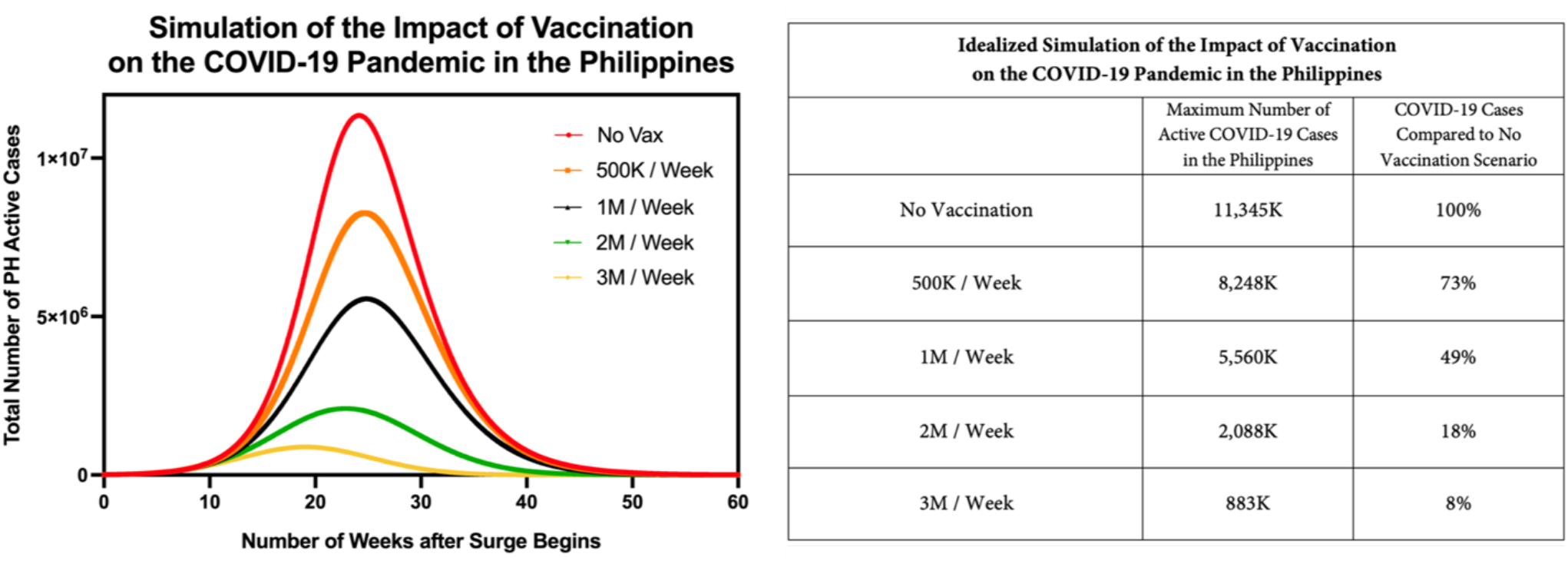
Idealized Simulation of the Impact of Vaccination on the COVID-19 Pandemic in the Philippines. Forecasted dynamics for the spread of the pandemic in the Philippines after the second surge of COVID-19 without any nonpharmaceutical interventions (NPI) like masking or social distancing in place. We modeled the impact of varying the rates of vaccination from 500K to 3M per week, equally distributed throughout the country.

**Figure 3:**
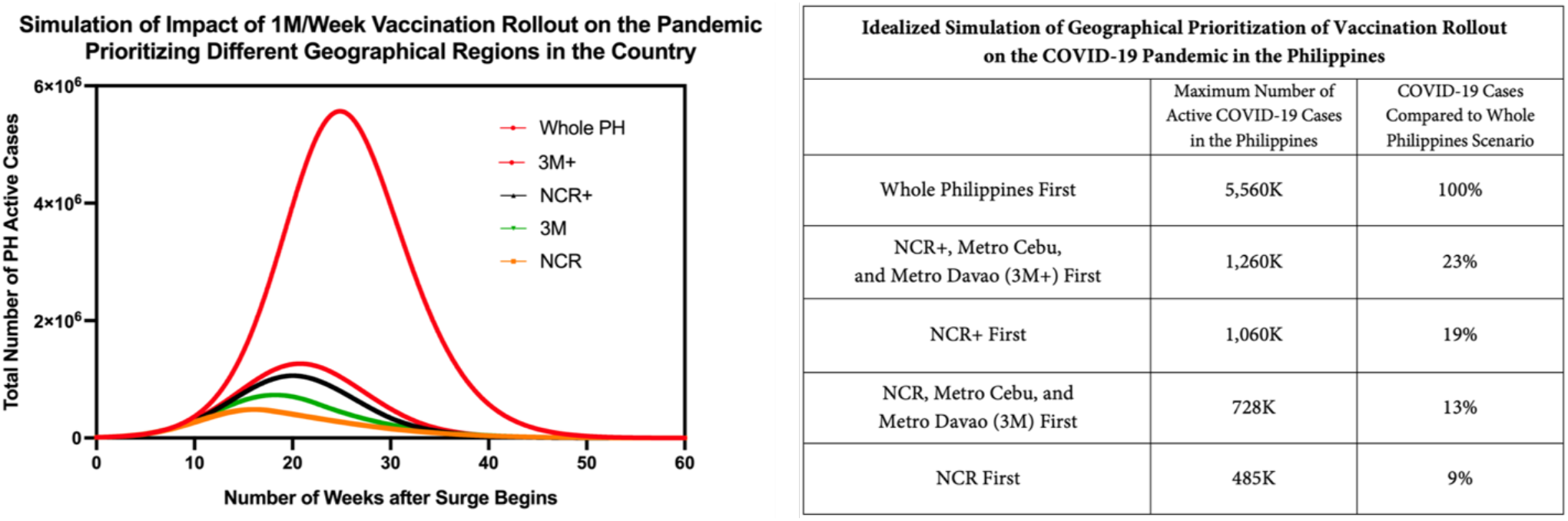
Idealized Simulation of the Impact of 1M/Week Vaccination Rollout on the COVID-19 Pandemic in the Philippines Prioritizing Different Regions in the Country. Forecasted dynamics for the spread of the pandemic in the Philippines after the second surge of COVID-19 without any nonpharmaceutical interventions (NPI) like masking or social distancing in place. We modeled the impact of prioritizing different regions of the country by deploying 1M vaccines per week to a particular region first before distributing them to the rest of the country.

We then turned to the primary question of interest for this study by comparing five vaccine deployment strategies that prioritize different geographical regions in the Philippines. As a baseline, we simulated a whole country rollout where vaccines would be equally distributed in the archipelago at a rate of 1M vaccinations per week. We then explored the effects of rolling out the vaccines at a rate of 1M per week in the NCR first (NCR), in Metro Manila, Metro Cebu, and Metro Davao first (3M), in the NCR Plus first (NCR+), in the NCR Plus, Metro Cebu, and Metro Davao first (3M+). Metro Manila, Metro Cebu, and Metro Davao are the three primary metropolitan regions of the Philippines. The NCR Plus is a geographical region encompassing the NCR and the adjacent provinces of Bulacan, Cavite, Laguna, and Rizal. This geographic “bubble” was placed under the highest level of quarantine to contain the second surge of COVID-19 in the capital region of the country. Note that our model is designed to transfer all the remaining vaccines to the rest of the country once the prioritized region has been fully vaccinated.

As shown in Figure 4, our simulation predicts that prioritizing the vaccine rollout in the NCR where all the vaccines would be sent to the capital would dramatically decrease the total number of COVID-19 cases in our idealized pandemic in the Philippines. By prioritizing the NCR, the national vaccine strategy would lead to a 91% decrease in the expected number of COVID-19 cases as compared to the whole country first scenario. This NCR first scenario (91% decrease in total cases) is better than either a Metro Manila, Metro Cebu and Metro Davao first scenario (87% decrease) or an NCR Plus first scenario (81% decrease). Therefore, our simulation suggests that the national vaccine strategy should target herd immunity in the NCR before inoculating the rest of the country in order to most efficiently halt the pandemic in the Philippines.

**Figure 4:**
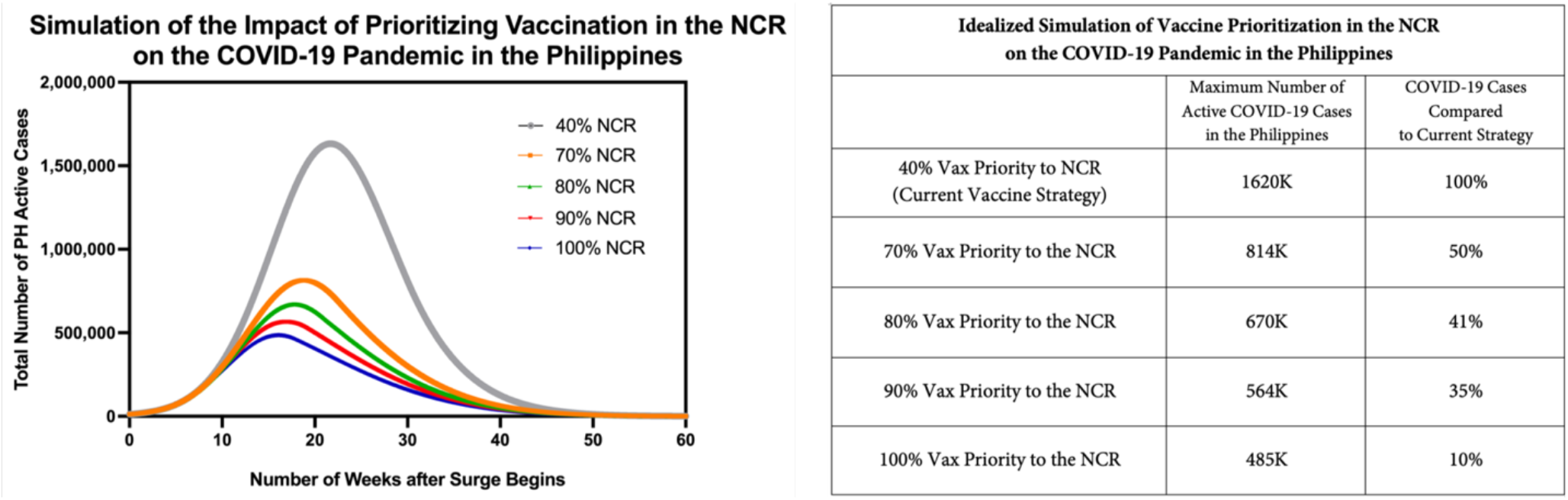
Idealized Simulation of the Impact of Prioritizing Vaccination in the NCR on the COVID-19 Pandemic in the Philippines. Forecasted dynamics for the spread of the pandemic in the Philippines after the second surge of COVID-19 without any nonpharmaceutical interventions (NPI) like masking or social distancing in place. We modeled the impact of varying the proportion of 1M vaccines deployed per week in the NCR. The remaining vaccines would be deployed to the rest of the country.

Finally, we wanted to explore the effect of varying the ratio of vaccines deployed to the NCR to vaccines sent to the rest of the Philippines. Numbers from the National Task Force for COVID-19 from May 11, 2021, revealed that the current vaccine deployment strategy of the national government distributes 40% of the national vaccine supply to the NCR (2,495,970 doses to the NCR out of 6,408,640 total doses distributed). As shown in Figure 5, our simulations reveal that increasing the percentage of vaccines deployed to the NCR from 40% to 100% would lower the number of total COVID-19 cases expected from the current strategy by 90%. However, we do not recommend committing the entire vaccine supply to the NCR since this strategy does not acknowledge that different segments of the population are at different risk for severe COVID-19 and death. Rather, we propose that the national government commit 90% of the vaccine supply to the NCR. The remaining 10% would be used to inoculate the medical frontliners and senior citizens who are most at risk throughout the country. Senior citizens (60 years old and above) make up about 9% of the total population of Filipinos according to published statistics (https://www.populationpyramid.net/philippines/). Our model revealed that deploying 90% rather than 100% of the vaccine supply to the NCR would still decrease the predicted COVID-19 case load from the current strategy by 65%. However, we would expect this strategy to decrease the total mortality rate since those most at risk throughout the archipelago would still be inoculated first.

Our study is limited by the simplicity of our 2SIR-VD model, which does not take into account the ever-changing non-pharmaceutical interventions that have been and are being implemented in the Philippines to mitigate the COVID-19 pandemic. However, its simplicity is also a strength because it allows us to account for the limitations and shortcomings of the epidemic data provided by the Department of Health of the Philippines. An earlier SIR model developed by one of us to model the pandemic in the NCR (Egwolf and Austriaco, 2020) was hampered by these shortcomings including changing definitions of what constituted a patient who had recovered from COVID-19, and inconsistent reporting schedules.

In sum, we have developed a 2SIR-VD model to determine if geographical prioritization of the national COVID-19 vaccine supply in the Philippines would have a significant impact on the dynamics of the pandemic. Our simulations reveal that prioritizing vaccine deployment to the National Capital Region (NCR) would significantly decrease the total case load of COVID-19 in the country. We therefore recommend deploying 90% of the available vaccine supply to the NCR to first achieve herd immunity there. The remaining 10% would allow the rest of the archipelago to vaccinate all of their senior citizens, thus shielding this vulnerable population against severe disease and death from COVID-19.

## Data Availability

No data sets were generated by this study.

